# Analyzing the Factor Structure and Sleep Quality of Pittsburgh Sleep Quality Index in Indian Information Technology Sector

**DOI:** 10.1101/2024.07.25.24308199

**Authors:** Arindam Chatterjee, Arijit Dutta, Rimu Chaudhuri

## Abstract

The Pittsburgh Sleep Quality Index (PSQI) has gained widespread acceptance as a useful tool to measure sleep quality. In order to formulate the diagnosis process, it is essential that we understand the factor structure inherent in the PSQI data. In this work, we seek to estimate such a structure with a focus on the Indian Information Technology (IT) workers. We have used Confirmatory Factor Analysis (CFA) and the Exploratory Factor Analysis (EFA) for this purpose. Instead of using random imputation to handle the frequently occurring missing values, we have used more sophisticated techniques thereby improving the accuracy of our results. We have also used the Multi layer perceptron based method to see how we can classify the sleep quality of the sampled population. We have discovered that, contrary to the general perception, most Indian IT employees have sleep quality belonging to good and very good classes.

**Author summary:** Arindam Chatterjee - Curently Assistant Professor of Computer Science and Engineering, Heritage Institute of Technology, Kolkata. Arindam has almost 16 years of teaching and research experience. In past he has worked in the industry for eight years. Area of academic interest is Impact of Information Technology on Indian Financial and Social sectors.

Rimu Chaudhuri -Currently Professor of Economics, Heritage Business School, Kolkata. Rimu is having teaching experience of almost 16yrs and research experience of 20 yrs. Area of academic / research interest is Entrepreneurship Development and for last couple of years Rimu is working in the field of Gender and Environment Sustainability. Last couple of years Rimu has worked extensively on Social Impact of IT. Rimu is also the President of Institution’s Innovation Council of Heritage Business School.

Arijit is a software engineer at Bloomberg LP, with a Master’s degree in Computer Science from the University of Florida. He specializes in creating and maintaining low-latency, high-performant, and robust backend applications. Arijit has a keen interest in Distributed Systems and Software Development. Currently, he contributes his expertise to the ESG (Environmental, Social, and Governance) sector, managing proprietary datasets and developing services that handle millions of requests per day. His work ensures efficient and reliable data management in a high-demand environment.

## Introduction

Information Technology in India is an industry consisting of two major components: IT services and business process outsourcing (BPO). The sector has increased its contribution to India’s GDP from 1.2% in 1998 to 7.7% in 2017. According to NASSCOM, the sector aggregated revenues of US $160 billion in 2017, with export revenue standing at US $99 billion and domestic revenue at US $48 billion, growing by over 13%. The United States accounts for two-thirds of India’s IT services exports [**?**]. The share of IT exports in the total exports revenue of Indian is around 30%.

A big part of the IT job outsourced from USA is BPO Support. For this the support person need to be available during the USA daytime hours. India being on the opposite side of the globe, has its night hours during the same time. So many IT support personnel in India have to stay awake at night or do night shifts to cater to American customers. Often the working hours are irregular, long and unplanned. All these pose a great problem to sleep quality of people working in the Indian IT sector. We guess most of these people working in the Indian IT sector may be having poor sleep quality.

Poor sleep quality has received increasing attention in various surveys and research. This is because poor sleep quality very often brings acute lifestyle problems. While sleep problems are widely studied in the West, there are very few studies in India. One possible reason could be due to a lack of valid and reliable measurement tools to assess sleep quality in such societies [1].

Since its introduction in 1989, the Pittsburgh Sleep Quality Index (PSQI) [2] has gained widespread acceptance. The inventory possesses good psychometric properties, including internal consistency, reliability, validity, as well as high correlations with sleep log data [1]. Because of this PSQI has been used in a variety of populations. In many cases PSQI has formed the basis of identifying responsible factors for sleep related disorders and hence became excellent enablers in the diagnosis process. Hence it is important to identify the basic factor based structure of PSQI results so that appropriate diagnosis and remedial measures can be adopted by specialists. The interesting observation here is that this factor based structure is not uniform across cultures and domains. This stems from the fact that sleep and its associated disorders have been shown to be quite dependent on the cultural background.

We have in [3] sought to understand the Factor Structure in a general Indian demographic context. The scope of the surveyed people were from a broad spectrum of the Indian society. In many works we have came across peculiar lifestyle adopted by Indian Information Technology (IT) workers. One of the main concerns reported by them was abnormal work schedule that was affecting their sleep and other health aspects. So we wanted to see how the PSQI based factor structure was applicable to the IT workers scenario in India. Below we seek to study this in detail.

### Literature Survey

Factor analysis uses mathematical procedures for the simplification of interrelated measures to discover patterns in a set of variables [4]. In statistics, confirmatory factor analysis (CFA) is a special form of factor analysis, most commonly used in social research. It is used to test whether measures of a construct are consistent with a researcher’s understanding of the nature of that construct (or factor). As such, the objective of confirmatory factor analysis is to test whether the data fit a hypothesized measurement model. In multivariate statistics, exploratory factor analysis (EFA) is a statistical method used to uncover the underlying structure of a relatively large set of variables. EFA is a technique within factor analysis whose overarching goal is to identify the underlying relationships between measured variables [5].

The single factor contains all the PSQI items. The 2-factor model has as the first factor the measurements of sleep efficiency which consists of the two PSQI items: sleep duration and habitual sleep efficiency. The second factor is sleep quality and consists of the remaining five items, viz., subjective sleep quality, sleep latency, sleep medication, sleep disturbance and daytime dysfunction. The 2-factor model is used from Magee’s [6]. The 3-factor model is as it is from Cole’s [7] and [6]. In this model first factor is sleep efficiency which consists of the two PSQI items sleep duration and habitual sleep efficiency as in 2-factor model. The second factor sleep quality consists of the three items: subjective sleep quality, sleep latency and sleep medication. The third factor daily disturbance is identified by the remaining two items sleep disturbance and daytime dysfunction. A notable feature of the factor analysis here is that an item is mapped to only one factor.

In [1], authors have discussed in detail about the various factor structure suggested and adopted by various researchers. The original developers of PSQI [2] recommended the use of a single factor. However most researchers have now found breaking down the PSQI into a multiple factor model can capture sleep quality better [1]. JC Cole et. el. has used Exploratory Factor Analysis (EFA) and Confirmatory Factor Analysis (CFA) approaches on a sample of elderly people to recommend the 3-factor [1]. Cole’s 3-factor was in a sample of PSQI study of 520 Nigerian university students. Magee et el has conducted the PSQI survey on a sample of 364 Australian adults and found both factor and 3-factor models give better results. In [8] authors have studied PSQI measures on 134 community dwelling renal transplant patients and found the 3-factor model better. Thus we see opinions differ about the most suitable factor structure drastically.

In [6] the authors have conducted the PSQI survey on a sample of 364 Australian adults and found both 2-factor and 3-factor models give better results. In [8] authors have studied PSQI measures on 134 community dwelling renal transplant patients and found the 3-factor model better. In [9] authors aim to validate the Italian version of the Pittsburgh Sleep Quality Index (PSQI), comparing five different groups of overall 50 individuals. The Italian version of the questionnaire provides a good and reliable differentiation between normal and pathological groups, with higher scores reported by people characterized by impaired objectively evaluated sleep quality.

In [10] authors aimed to assess the reliability, validity, and factor structure of the Persian version of PSQI. They conducted a cross-sectional survey on 1115 citizens of Arak City,Iran aged 18-60 years, selecting candidates by stratified random sampling method. They used EFA with a 2-factor structure, following it up with CFA to assess the reliability of PSQI questionnaire. Their findings suggest that the Persian version of PSQI displayed satisfactory validity and reliability to measure the quality of sleep of Iranian people. Thus we see opinions differ about the most suitable factor structure drastically.There is however a universal agreement on the reliability and validity of PSQI, verified in some cases with clinical tests as mentioned above.

Authors in [11] aim to investigate the dimensionality of the underlying factor structure of the PSQI in a multi-ethnic working population in Singapore. The PSQI was administered to full-time employees participating in a workplace cohort study. They used EFA followed by CFA and identified a two-factor model with reliable internal consistency and goodness-of-fit. They recommend using the two-factor model to assess sleep characteristics in working populations in Singapore, given that it performs comparable to the three-factor model and is simpler compared to the latter.

### Indian Context for PSQI

Unfortunately not a whole lot of focus has been given on the PSQI in Indian context so far. In [12], authors present the study in an Indian university student context, a common ground for sleep disorder related problems. Little effort has been spent to discover the influence of latent factors in the sample however. In [13] authors survey more than 1000 healthy people in South India using PSQI and other suitable measures of sleep related disorders (SRD). In our previous work [3], we sought to explore the factor structure from the PSQI index using surveyed data from a broad spectrum of the Indian society.

In our present we seek to overcome some of these deficiencies. Here we specifically target the Indian IT sector where because of peculiar work and lifestyle, people tend to suffer from sleep related problems. We evaluate via CFA the appropriate factor structure suitable in this context in the line as in [1]. One deficiency of this survey is that since the majority of the IT workers are in the young age group, we could not conduct a proper analysis across various age groups but had to resort to other available factor diversity like gender and medication etc.

## Materials and methods

### Conduction of the survey

PSQI is a self-rating tool designed to assess a wide variety of indicators of sleep quality during the month before testing. It consists of 19 items from which 7 component scores are calculated and summed into a global score; they are subjective sleep quality, sleep latency, sleep duration, habitual sleep efficiency, sleep disturbances, use of sleep medication, and daytime dysfunction. The score of each component has a range of 0 to 3, yielding a global score of sleep quality ranging from 0 to 21, with a higher score indicating poorer sleep quality ([1], [2]). Although it is a practice of assigning values 0 to 3 for each of the components in the questionnaire, we chose 1 to 4 as the range (to avoid problems arising out of use of 0 in certain cases). Also we inverted the interpretation of the scale, i.e., we chose a higher value to represent better sleep quality. [This is opposite to what is reported in [1], [2] etc.].

As mentioned before, we chose respondents randomly. They were all known to the authors, with various gender and other parameters. One of the common criterion was that all were somehow related to the Indian IT industry by profession. As we said before, the age group lacked diversity as there were hardly anybody working in the IT industry within the age group 40+ or more. So in order to bring in more meaningful age related diversity in the population, we included people from other spectrum as well most. Obviously people representing higher age groups were not even thinly connected to the Indian IT sector. We deviated from [1] from the age groups as many respondents were from the students community where the authors have more access. Remaining respondents were from the neighbours, relatives and friends circle. In all we could collect a sample of 252 respondents. The grouping details are shown in Table 1.

**Table 1.** PERSONAL CHARACTERISTICS OF PARTICIPANTS (N= 252)

|  | Age 18-19 | Age 30+ |
| --- | --- | --- |
| Male | 136 | 34 |
| Female | 66 | 16 |
| Total | 202 | 50 |

**Table 2.** GOODNESS OF FIT OF OUR MODELS (N=252))

| Model | Number of factors | Age | SRMR | RMSEA | CFI |
| --- | --- | --- | --- | --- | --- |
| 1 | 1 | All | 0.076 | 0.127 | 0.766 |
| 2 | 2 | All | 0.063 | 0.104 | 0.854 |
| 3 | 3 | All | 0.056 | 0.096 | 0.898 |
| 4 | 1 | 30+ | 0.085 | 0.104 | 0.908 |
| 5 | 2 | 30+ | 0.083 | 0.104 | 0.914 |
| 6 | 3 | 30+ | 0.083 | 0.125 | 0.896 |
| 7 | 1 | < 30 | 0.085 | 0.144 | 0.700 |
| 8 | 2 | < 30 | 0.070 | 0.113 | 0.828 |
| 9 | 3 | < 30 | 0.062 | 0.105 | 0.874 |

Please note that since there were a large number of students present in the sample, we made a new group (compared to [1]) for the age group 18-29. This is consistent with results in Indian context where students and young people exhibit a lot of sleep related disorder problem. Also the number of 60+ people was small, so we did not create any subgroups among them (in [1] there are two subgroups created, one for 60-74 years, and the other for 75+ years of age). In fact the sample size in the age group 30-59 was also so small compared to the 18-29 group that we chose to combine all into a single group 30+. This reduces the group count by one compared to our earlier work in [3].

The level of education of the respondents was obviously high as IT sector requires personnel with some level of higher education as their background. This eliminated the need of trained interviewers and made the process fast and simple. We think this actually reduced the confusion inherent in such surveys and zeroed the probability of wrong answers. The modes of the interview were response to printed sheets, email replies and, phone and face to face conversations. The survey remained anonymous.

### Factor analysis of the survey data

In statistics, Confirmatory Factor Analysis (CFA) is a special form of factor analysis, most commonly used in social research. It is used to test whether measures of a construct are consistent with a researcher’s understanding of the nature of that construct (or factor). As such, the objective of confirmatory factor analysis is to test whether the data fit a hypothesized measurement model. Both EFA (Exploratory Factor Analysis) and CFA are employed to understand shared variance of measured variables that is believed to be attributable to a factor or latent construct. Despite this similarity, however, EFA and CFA are conceptually and statistically distinct analyses. The goal of EFA is to identify factors based on data and to maximize the amount of variance explained. By contrast, CFA evaluates a priori hypotheses and is largely driven by theory. EFA is often considered to be more appropriate than CFA in the early stages of scale development because CFA does not show how well your items load on the non-hypothesized factors. Another strong argument for the initial use of EFA, is that the misspecification of the number of factors at an early stage of scale development will typically not be detected by CFA. At later stages of scale development, CFA may provide more information by the explicit contrast of competing factor structures.

In CFA, several statistical tests are used to determine how well the model fits to the data. Kline (2010) recommends reporting the Chi-squared test, the Root mean square error of approximation (RMSEA), the comparative fit index (CFI), and the standardized root mean square residual (SRMR) [14]. The chi-squared test indicates the difference between observed and expected covariance matrices. Values closer to zero indicate a better fit. One difficulty with the chi-squared test of model fit, however, is that researchers may fail to reject an inappropriate model in small sample sizes and reject an appropriate model in large sample sizes. The root mean square error of approximation (RMSEA) avoids issues of sample size by analyzing the discrepancy between the hypothesized model, with optimally chosen parameter estimates, and the population covariance matrix. The RMSEA ranges from 0 to 1, with smaller values indicating better model fit. Standardized root mean square residual (SRMR) is the square root of the discrepancy between the sample covariance matrix and the model covariance matrix. The goodness of fit index (GFI) is a measure of fit between the hypothesized model and the observed covariance matrix. Comparative fit indices (CFI) compare the chi-square for the hypothesized model to one from a “null”, or “baseline” model. This null model almost always contains a model in which all of the variables are uncorrelated, and as a result, has a very large chi-square (indicating poor fit).

CFA was conducted in our work using the “lavaan” package [**?**] with “R” software. “lavaan” is a package for structural equation modeling implemented in the “R” system for statistical computing. “lavaan” is anacronym for latent variable analysis, and its name reveals the long-term goal: to provide a collection of tools that can be used to explore, estimate, and understand a wide family of latent variable models, including factor analysis, structural equation, longitudinal, multilevel,latent class, item response, and missing data models [15]. The choice, composition and nature of factors used for various test runs is similar to what is proposed in existing literature and is already described earlier in the section.

In addition to using data from all participants, CFA employed data from different age groups, one aged 18 to 29 (the young ones), another aged 30 and above (the young, the middle aged and the old). The breakup is somewhat different from other sources for two reasons. First, we had samples with age values varying very widely (smallest 18 to maximum 89). But since we were focusing mostly on Indian IT employees with an average age only in the twenties, we have a very high concentration of data at that age group. Second, since the number of surveyed population was small, having age groups as discussed in other references ranging over smaller intervals, take [1] for example, would have yielded an insignificant number of samples per group. Unfortunately CFA would not be able to converge to a solution with such a small sample size per group. Hence our choice. One problem with our group selection is that some groups will not be able to identify the group characteristics very well. Obviously a 30 something person is grossly different from a person of 55. This is a deficiency we have identified.

Furthermore, CFA fitted multiple age group data by constraining factor loadings as equal across the groups, as well as allowing factor loadings to be different among the groups. As can be found in [16], the “R” package has a module that can simultaneously compare models with free loading as well as different levels of constrained factor loadings. We only compared the first two of these models as stated above.

To have a common metric across the three age groups, the CFA used standardized scores of the seven components as inputs. Altogether, this analyses involved nine models for different numbers of factors and different sets of data (see Table 3). A model demonstrated a good fit to the data when its standardized root-mean square residual (SRMR) was below 0.05, root-mean square error of approximation (RMSEA) below 0.07, and Comparative Goodness-of-Fit Index (CFI) above 0.95 [17].

**Table 3.** 2-FACTOR FACTOR LOADINGS STANDARDIZED FOR THE WHOLE SAMPLE (N = 252)

| Indicator | Model 2 | Model 5 | Model 8 |
| --- | --- | --- | --- |
| Sleep efficiency |  |  |  |
| Sleep duration (C3) | 1.0 | 1.0 | 1.0 |
| Habitual sleep efficiency (C4) | 1.002 | 0.893 | 0.990 |
| Sleep quality |  |  |  |
| Subjective sleep quality (C1) | 1.0 | 1.0 | 1.0 |
| Sleep latency (C2) | 1.750 | 3.494 | 1.518 |
| Sleep medication (C6) | 0.593 | 2.344 | 0.233 |
| Sleep disturbances (C5) | 0.893 | 1.906 | 0.741 |
| Daytime dysfunction (C7) | 0.702 | 1.417 | 0.544 |

### Handling Missing Values

Missing data are a key problem in statistical practice. Most statistical methods cannot be applied directly on an incomplete dataset. One of the common approaches to deal with missing values consists in imputing missing values by plausible values. It leads to a complete dataset that can be analyzed by any statistical method. However, the interpretation of the results should be done with caution, since there is necessarily uncertainty associated with the prediction of values [18].

If values are missing completely at random, the data sample is likely still representative of the population. But if the values are missing systematically, analysis may be biased. Values in a data set are missing completely at random (MCAR) if the events that lead to any particular data-item being missing are independent both of observable variables and of unobservable parameters of interest, and occur entirely at random. When data are MCAR, the analysis performed on the data is unbiased; however, data are rarely MCAR. Missing at random (MAR) occurs when the missingness is not random, but where missingness can be fully accounted for by variables where there is complete information. An example is that males are less likely to fill in a depression survey but this has nothing to do with their level of depression, after accounting for maleness. Missing not at random (MNAR) (also known as nonignorable nonresponse) is data that is neither MAR nor MCAR (i.e. the value of the variable that’s missing is related to the reason it’s missing). In the previous example, if men failed to fill in a depression survey because of their level of depression, that would be MNAR type.

In [19], Bayesian methods for missing data are reviewed from the Calibrated Bayesian (CB) perspective. The basic theory of the Bayesian approach, and the closely related technique of multiple imputation, is described. Then applications of the Bayesian approach to normal models are described, both for monotone and nonmonotone missing data patterns.

Most of the data in PSQI is categorical data (or ordinal data) apart for the ones related to sleep latency and sleep duration. Most of the missing values occur very randomly here. So we tend to think that this is the MCAR case. But on closer observation we found that quite a few missing values belonged to the Female respondents. So we think there are some indications of MAR type missing also. The amount of missing information is less than 10%. If there is one missing value for a particular respondent, chances of more missing pieces increase.

The imputation of categorical data can be done with non-parametric methods such as the K-nearest neighbours or with parametric model methods. The most classical model of these is the log-linear one. It has a major drawback: the number of parameters increases rapidly with the number of levels of categorical variables. Therefore, in practice, it becomes unusable in some cases. Other models have been proposed to overcome this problem, such as the latent class model, or the loglinear hierarchical model [18].

In [18], authors propose a new method to handle the imputation (for mixed data however). This is based on the Principal Components method and appropriately named as Factorial Analysis for Mixed Data (FAMD) (also known as PCAMIX). In this work we have used the random imputation method to get the initial results. In the lavaan package in R facilitating the CFA and EFA methods, the default option for the default model (i.e. ML, maximum likelihood) to handle missing values is listwise deletion, i.e., discount rows with missing values. Since there are a substantial percentage of missing values in our survey data, we think this sort of direct deletion of data can give inaccuracy. So instead we are using the inbuilt missing value method “fiml” indicating Full Information Maximum Likelihood method.

FIML is widely reported in literature as the most appropriate method to adopt to handling missing values when it comes to MCAR or MAR types.

In [20], authors used a A Monte Carlo simulation to examine the performance of four missing data methods in structural equation models: full information maximum likelihood (FIML), listwise deletion, pairwise deletion, and similar response pattern imputation. The effects of three independent variables were examined (factor loading magnitude, sample size, and missing data rate) on four outcome measures: convergence failures, parameter estimate bias, parameter estimate efficiency, and model goodness of fit. Results indicated that FIML estimation was superior across all conditions of the design. Under ignorable missing data conditions (missing completely at random and missing at random), FIML estimates were unbiased and more efficient than the other methods. In addition, FIML yielded the lowest proportion of convergence failures and provided near-optimal Type 1 error rates across both simulations.

FIML has been shown to be an effective way of handling missing values in clinical conditions similar to sleep related disturbance. In [21] authors have analyzed the type of missing values that frequently occur in Alcohol Clinical Trials. They observed that the rate of participant attrition in these trials is often substantial and can cause significant issues with regard to the handling of missing data in statistical analyses of treatment effects. It is common for researchers to assume that missing data is indicative of participant relapse and under that assumption many researchers have relied on setting all missing values to the worst case scenario for the outcome. This sort of single imputation method has been criticized for producing biased results in other areas of clinical research as well. FIML produced the least biased effect estimates and standard errors in such clinical missing value scenarios, compared to other techniques that definitely proved to be very much biased and producing unacceptable results. This situation seems contextual in PSQI observations also.

### Classification using MLP

Multi-Layer Perceptron(MLP) is a type of artificial neural network which is a powerful tool used for classification. It is a type of surpervised learning model. A perceptron is a single neuron model that was a precursor to larger neural networks. Each node, apart from the input nodes, has a nonlinear activation function. MLP uses backpropagation as a supervised learning technique. The model is trained using the data that is present. Then it is tested with respect to other data whose outputs are known. The accuracy can then be calculated from the result.

We have used python programming language to implement the MLP model. The data collected in the survey were used to train and test the model. Our model consisted of 7 inputs (C1 to C7 components). Apart from the 7 components, two more columns were computed. They were the Global PSQI ‘gpsqi’ and ‘score’. The ‘gpsqi’ column is the sum of all the seven components, that is used to rate the sleep quality of a person. Each component varies from 1 to 4 (1 being worst and 4 being best) and thus ‘gpsqi’ can vary between 7 to 28 (7 denoting the worst sleep quality and 28 denoting best sleep quality of a person). For the purpose of classification we have divided ‘gpsqi’ into 4 classes which is stored in the ‘score’ column and which acts as the output of out MLP. People with ‘gpsqi’ less than 7 are given the class ‘1’ (very bad sleep quality class) in ‘score’ column, between ‘7’ and ‘14’ are given class ‘2’(bad sleep quality class), between ‘14’ and ‘21’ are given class ‘3’(good sleep quality class) and greater than ‘21’ are given class ‘4’(very good sleep quality class).

## Results

### Participant characteristics

Among the 252 candidates, we had more representations from the age group 18-29 for the reason cited before (since IT employees, focus of our survey, are mostly young).

### Confirmatory factor analysis

From the Table 2, we note that for all data present, we see gradual improvement in the fitness values form 1-factor to 3-factor models (Models 1, 2, 3). The CFI values increase from 0.766 to 0.898. The SRMR values show similar patterns (0.076 to 0.056). Same for the RMSEA values. So we conclude that the 3-factor model is more suited.

For the 30+ age group data (corresponding models are 4,5,6), however the 2-factor model shows better fit (CFI = 0.914) than the 1-factor model (0.908) and the 3-factor model (0.896). For the target group of our work here, i.e., Indian IT workers, most of whom belong to the 18-29 age group, the 3-factor model looks the best.

Thus we conclude that the sample data indicates towards adopting a two factor model for PSQI (Model 5) for 30+ age group. For the age group <30, the 3-factor model (Model 9) looks better; the same is also true for all age group (Model 3) where the 3-factor model shows the best goodness of fit. But all of these fall below the required fitness criteria laid down earlier (0.95).

We see from Table 3 that Model 5 where the two factor model is used, factor loadings are quite interesting across the models. For the age group 30+ (Model 5), all the indicator variables are loaded heavily on corresponding factors. In particular the indicator variables Sleep Latency, Disturbance and the Sleep Efficiency show deep impact on corresponding factors. For Model 2 (for all group data), we see somewhat similar results – all components are heavily loading corresponding factors.

For the 3-factor loadings, from Table 4, for models 3 and 9, we see similar pattern, all components are heavily loading corresponding factors. However C4 (habitual sleep efficiency) and latency (C2) are loading with highest values.

**Table 4.**
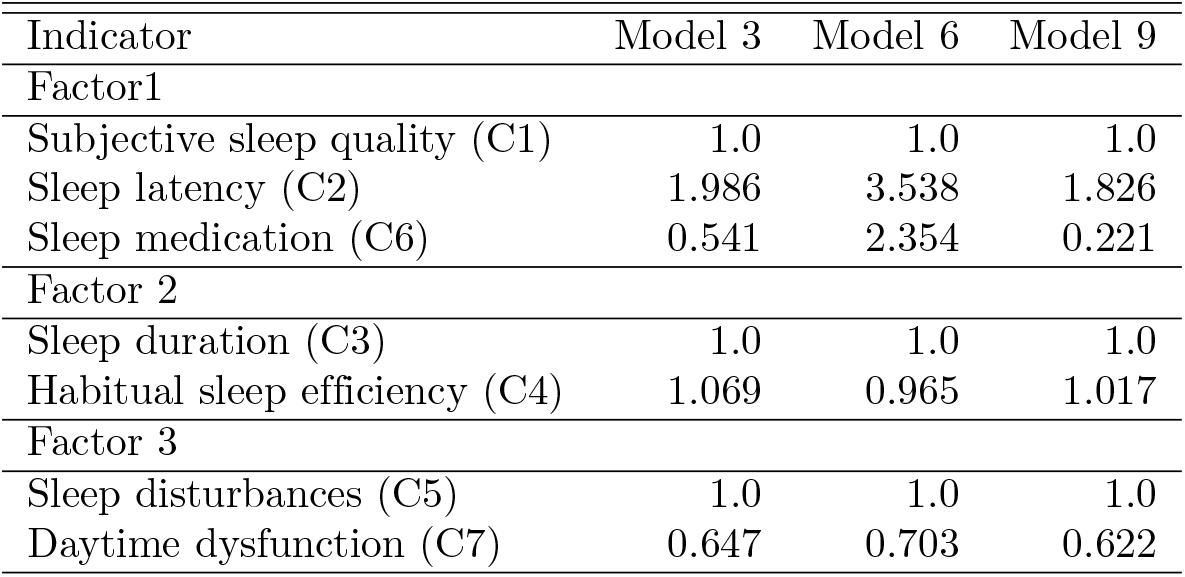
3-FACTOR FACTOR LOADINGS STANDARDIZED FOR THE WHOLE SAMPLE (N = 252)

### Exploratory factor analysis

Since it is obvious from the discussion above that CFA using the factor structure explored in existing literature does not give very good results, it is better to resort to own Exploratory Factor Analysis (EFA). We used the “factanal” method available inside “R” to find out the latent factor structure available inside our data. In order to complete the factorization process, we had used two most popular types of rotation tactics: “varimax” (the default one) and “promax”. It is no wonder that we found the factor structure quite different from that proposed in literature. This structure is shown in Table 5. We have noted p-values corresponding to these models (not shown here). In all cases, the 3-factor model has significant values whereas the 2-factor model does not, indicating the 3-factor model is more suitable. We also observe that the 3-factor model in both cases have the same factor structure. The 2 factor model for varimax is exactly same as the CFA 2-factor model originally proposed.

**Table 5.**
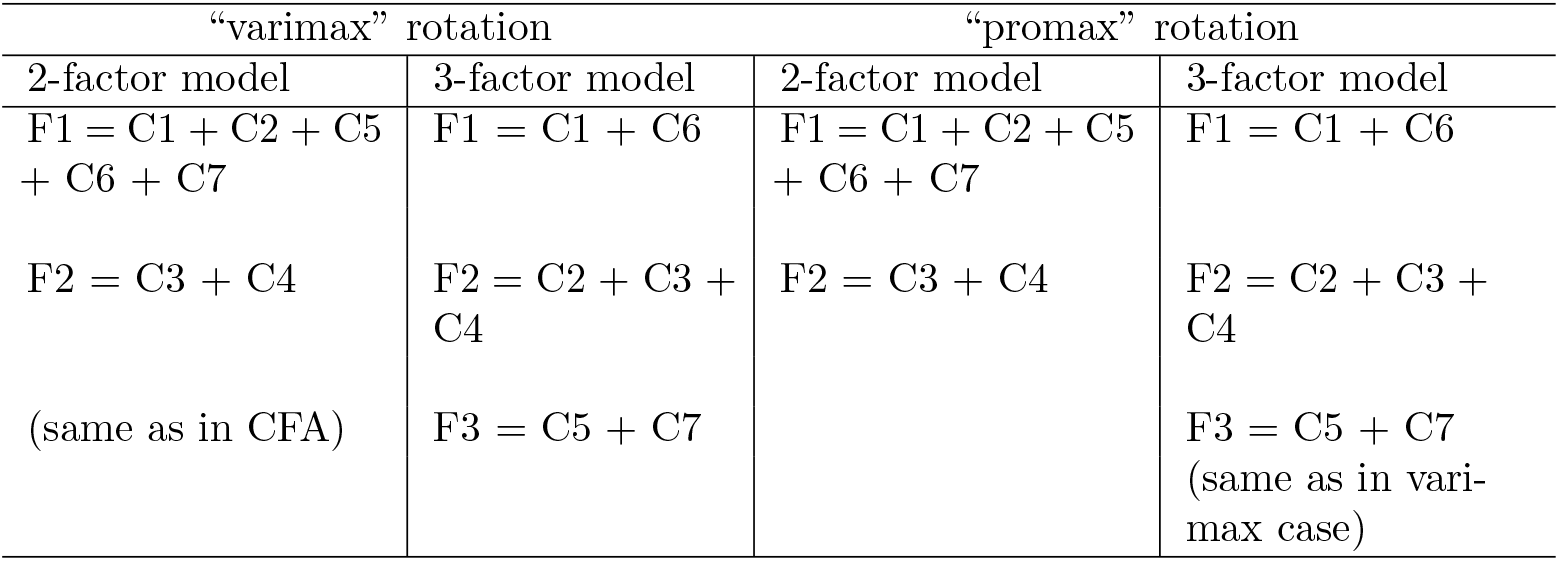
FACTORS AFTER EXPLORATORY FACTOR ANALYSIS.

After deciding upon the various factors, we ran CFA on the various datasets mentioned above. We have tabulated the results in Table 6. For model 2, for promax, there is no improvement. For model 3 also there is no improvement. So the original CFA is better for all age group data. For model 5, we see the original result and our result are similar (CFI = 0.914). However for model 6, the 3-factor model, our model is much better (0.969) than the original model (0.896). This is a remarkable feat esp that the original CFA predicted the suitability of a 2-factor model whereas our result reveals adopting a 3-factor model is a better idea.

**Table 6.** GOODNESS OF FIT OF OUR MODELS (N=252) (OUR EFA)

| Model | # factors | # groups | Age | SRMR | RMSEA | CFI |
| --- | --- | --- | --- | --- | --- | --- |
| 2 | 2 (varimax) | 1 | All | Same as in | CFA Model 2 |  |
| 2 | 2 (promax) | 1 | All | 0.063 | 0.104 | 0.854 |
| 3 | 3 (vari-/promax) | 1 | All | 0.058 | 0.111 | 0.860 |
| 5 | 2 (varimax) | 1 | 30+ | Same as in | CFA Model 5 |  |
| 5 | 2 (promax) | 1 | 30+ | 0.083 | 0.104 | 0.914 |
| 6 | 3 (vari-/promax) | 1 | 30+ | 0.076 | 0.068 | 0.969 |
| 8 | 2 (varimax) | 1 | < 30 | Same as in | CFA Model 8 |  |
| 8 | 2 (promax) | 1 | < 30 | 0.070 | 0.113 | 0.828 |
| 9 | 3 (vari-/promax) | 1 | < 30 | 0.072 | 0.140 | 0.775 |

For model 8, the original solution was similar to ours. For model 9, the original approach gave a much better result however (0.874 vs 0.775). Also in the new structure, the 2-factor model looks better whereas in the original model 3-factor analysis looks better. So we suggest adopting the 3-factor model in case of the age group < 30 years.

### Results from handling Missing Values

We note from Table 7 that for the age group 30+, the fitness value improves mostly with the FIML method of handling missing values. For other age groups the change is not giving any improvement. We think this may because most of the missing values occurred in the 30+ age group than the <30 group.

**Table 7.** CFA PERFORMANCE COMPARISON OF VARIOUS MISSING VALUE HANDLING TECHNIQUES.

| Imputation<br>Technique | Random | Imputation |  | Inbuilt | in R | (“fiml”) |
| --- | --- | --- | --- | --- | --- | --- |
| Model | SRMR | RMSEA | CFI | SRMR | RMSEA | CFI |
| 1 | 0.076 | 0.127 | 0.766 | 0.073 | 0.104 | 0.758 |
| 2 | 0.063 | 0.104 | 0.854 | 0.057 | 0.109 | 0.863 |
| 3 | 0.056 | 0.095 | 0.898 | 0.049 | 0.096 | 0.910 |
| 4 | 0.085 | 0.104 | 0.908 | 0.073 | 0.123 | 0.898 |
| 5 | 0.083 | 0.104 | 0.914 | 0.060 | 0.075 | 0.965 |
| 6 | 0.083 | 0.125 | 0.896 | 0.059 | 0.084 | 0.962 |
| 7 | 0.085 | 0.144 | 0.700 | 0.082 | 0.155 | 0.694 |
| 8 | 0.070 | 0.113 | 0.828 | 0.066 | 0.125 | 0.816 |
| 9 | 0.062 | 0.105 | 0.874 | 0.058 | 0.120 | 0.856 |

### Results from the MLP Classification

The description of the ‘score’ column is shown in Fig 1. We could not find any data belonging to the class ‘1’ sleep quality thus is absent in the below picture. We can also see that we have only 4 data that belongs to class ‘2’. Thus signifying that most of our data belongs to good ‘3’ and very good ‘4’ classes.

**Fig 1.**
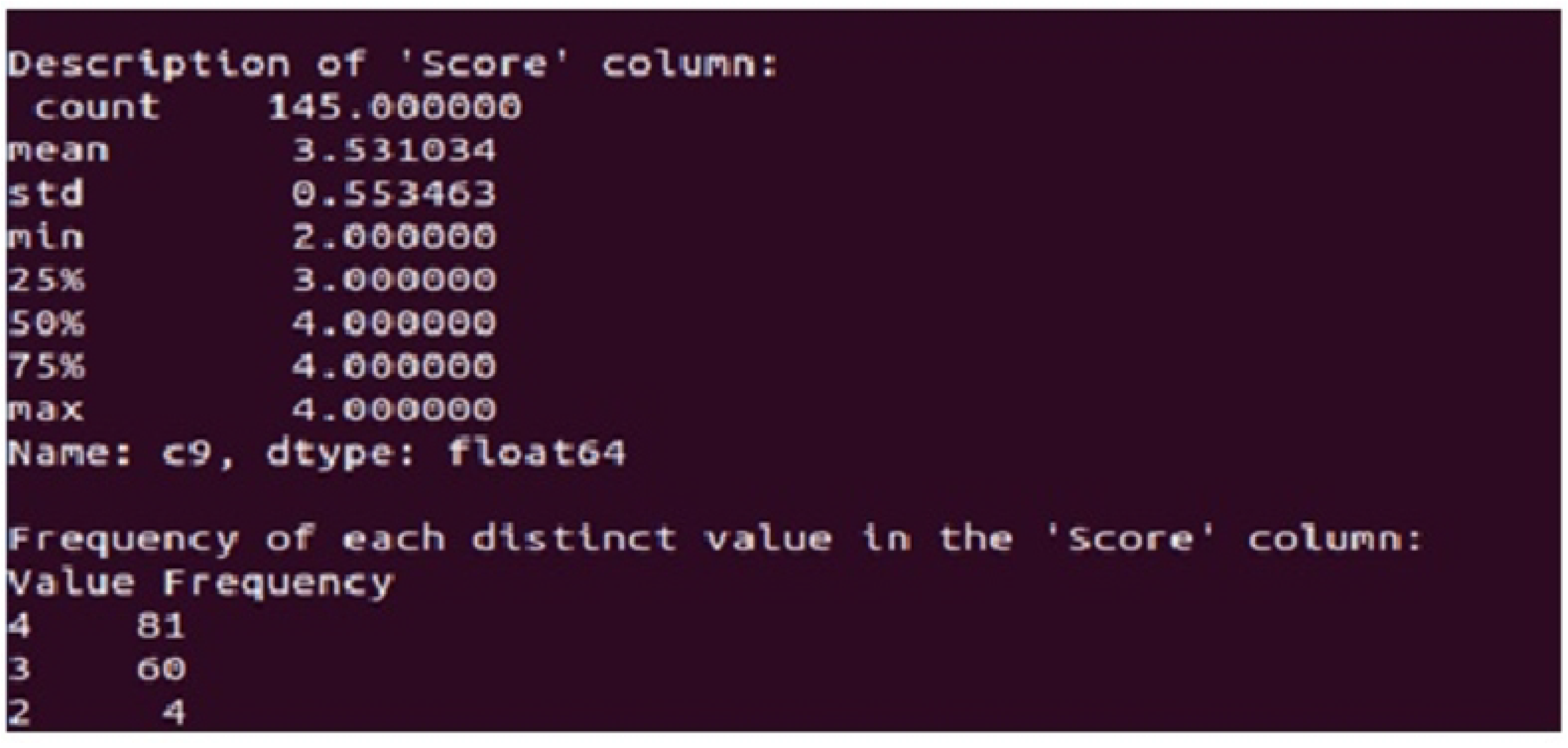
DESCRIPTION OF THE MLP SCORE COLUMN.

The ‘score’ column was used as the basis of judging the overall sleep quality class of a person and so was our output. The data was split into training and testing sets. 65 % of the data was used for training and the rest 35 % was used for testing. 3 hidden layers were used with 7 neurons in each layer. The output of the MLP model would be either ‘1’, ‘2’, ‘3’ or ‘4’ depending on the 7 components. The model would learn using the 65% of training data and then use the 35% of the test data to predict the sleep quality class(output). The output class thus obtained from the model would be compared to the output class that has been already calculated and the accuracy of the model would be checked.

A confusion matrix is a summary of prediction results on a classification problem. The number of correct and incorrect predictions are summarized in Fig 2 with count values and broken down by each class. The accuracy of the model is 96%.

**Fig 2.**
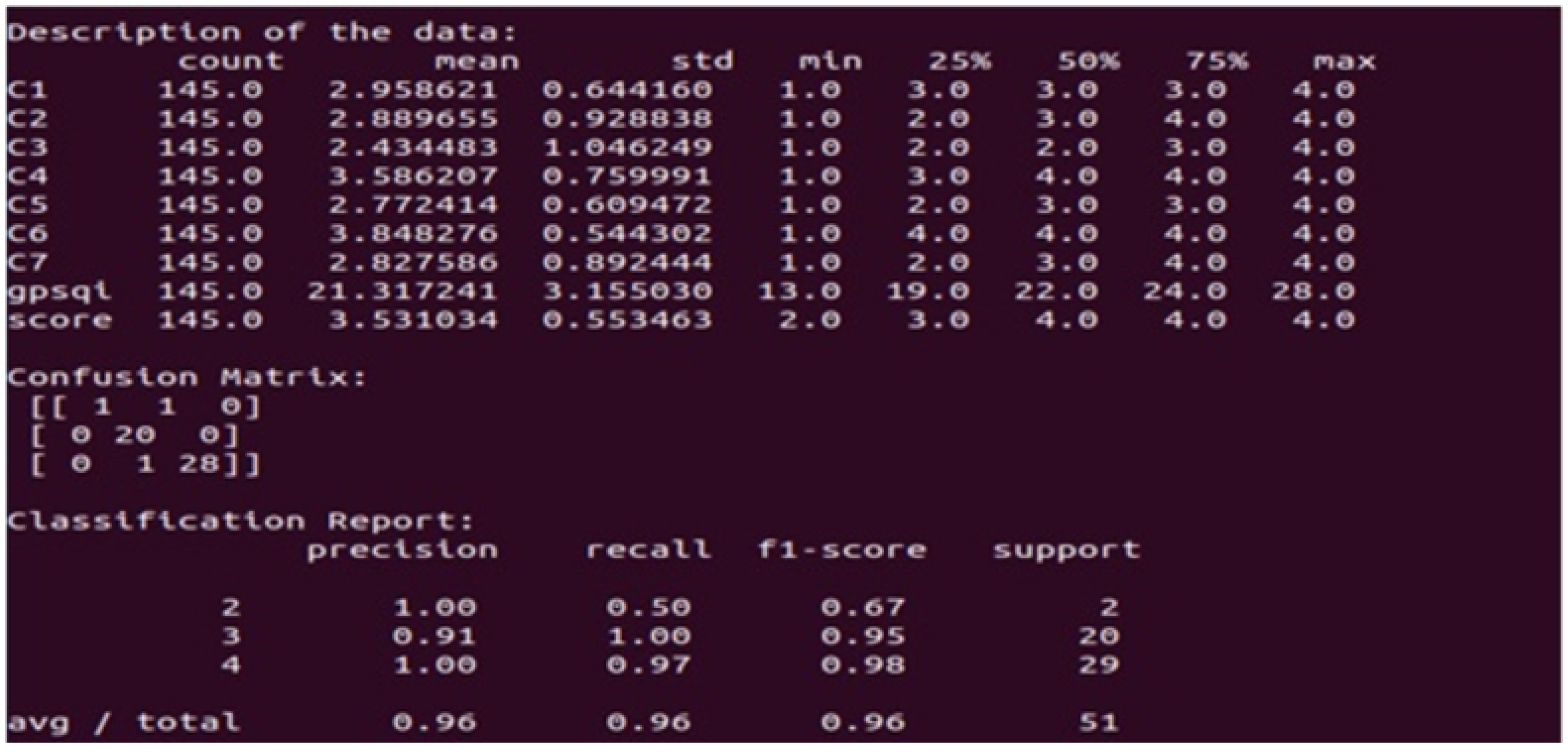
DESCRIPTION OF THE MLP DATA AND CLASSIFICATION.

## Discussion

In this work we conducted a pilot study of 252 Indian males and females belonging to various age groups to find out the sleep quality prevailing among them. Our main aim was to find the sleep quality and the corresponding PSQI factor structure among the young IT workers of India. We employed the CFA methods. We used initially the 1-, 2-and 3-factor structures proposed in existing literature. One key characteristics of this breakup was that one indicator variable only loaded one factor. We found that the two factor and the three factor models better fit in some cases whereas the 1-factor model did not fit as well. This is in line what is reported in our earlier work [3] and the Cantonese version of the report [1].

Our work however differs from existing reports like [1] in many aspects. First of all we see far less sleep problem reported by Indians (27%) compared to Westerners (40%-50%) and Cantonese people (58%). Second, we see there is no particular fit of factors for some cases. Third, we see a strong correlation between various factors related to sleep quality that was not reported in other places. One of them is Sleep Medication. For the first case we attribute the difference related to cultural reasons. It is generally accepted that sleep related problems can be attributed to cultural heritage. In [1], Cantonese old people are shown to be more prone to sleep related disorders as they had undergone terrible socio political unrest in their lifetime. In Indian context such legacy may be absent.

In order to address the second issue we deviated from the existing factor structure. We created a new factor structure based on our survey data using EFA. We found the new structure substantially different as is evident from the fact that some single indicator variables loaded more than one factor (not shown in the final model structure however as their loading was small). Using these models we ran CFA, and we found the CFI measures improving in many cases. In some cases however we still did not find good fits. This may be because of a small dataset.

For the third case, we explored in particular what is the sleep quality of the Indian IT sector employees who formed the bulk of the <30 years age group. We found that the sleep quality of this age groups is quite good, contrary to the general perception held about them. We however strongly believe the perception is true and what we have observed is just a sample bias. Most of our surveyed population was from the fresher group (22-24 years age group). This group may be less affected by sleep related discrepancy. However we should have more experienced IT people to draw more accurate conclusion on this.

The present study however suffers from a number of limitations. First of all, as repeatedly mentioned, the number of participant was too small to carry out CFA so that its results can be deemed very convincing. We did not include the attribute Marital status in the sample. This is included in [1] and we suspect has some relevance to sleep disorder. Also a wider group of participants with lesser level of education should be included. This is not possible with the IT workers. In [1] it is stated that the level of education especially among the elderly play an important role in determining sleep quality. This may be attributed to the more socio economic pressure this group of people has to undergo. Also as we have stated earlier we have group structures having very broad range of values. Hence some groups may not be representing their population in a best possible manner.

## Conclusion

We have conducted a pilot study on 252 Indian males and females belonging to different age groups regarding their sleep quality. The popular and powerful PSQI questionnaires were used to identify indicator variables necessary for producing good sleep quality. We found that Indians in general have less sleep disturbance compared to Westerners and Cantonese people. In particular Indian IT people show good sleep quality. This is contrary to the general perception. We however attribute this to the sample bias. Since identifying latent factors responsible sleep quality can help diagnose sleep problems for medical usage, we employed CFA to identify responsible factors. Using existing literature, 1-, 2- and 3-factor models were used. We found that 1-factor model gives a poor fit measure. The 2-factor and 3-factor models give good measures in some cases (across various age group data) but fail to either give a good fit in other cases.

Hence we employed EFA to identify responsible factors on our own. We discovered quite different latent factor structure. Using two different popular rotation methods (varimax and promax) we came up with new CFA measures. We have seen in certain cases where the original CFA failed to find a good solution, our model fitted better. However in other cases it failed. We suspect this anomaly owing to small dataset size. This however needs further investigation.

## Data Availability

Data cannot be shared publicly because of privacy reasons. But if required can be shared with some limitations.

## Acknowledgments

The authors like to acknowledge all their coworkers in various IT companies in India and students at Heritage Institute of Technology, Kolkata for enthusiastically taking part in the PSQI survey.

## References

1. Ohno S. Alice M., Chong L., Cheung Chau. Factor Structure of a Cantonese-version Pittsburgh Sleep Qualilty Index. Sleep and Biological Rhythms 2016.

2. D.J. Buysse D.J.,Reynolds C.F., Monk T.H., others. The Pittsburgh Sleep Quality Index: a New Instrument for Psychiatric Practice and Research. Psychiatry Research 1989;193–213.

3. Dutta Arijit, Chatterjee Arindam. A Pilot Study Examining the Factor Structure of Pittsburgh Sleep Quality Index in Indian Context. Grenze International Journal of Engineering and Technology 2019;55-62.

4. Child D. The Essentials of Factor Analysis (3rd Ed.). Continuum International Publishing Group. New York, New York.; 2006.

5. Norris Megan, Lecavalier Luc. Evaluating the Use of Exploratory Factor Analysis in Developmental Disability Psychological Research. Journal of Autism and Developmental Disorders 2009;8-20.

6. Magee C.A.,Caputi P., Iverson D.C., others. An Investigation of the Dimensionality of the Pittsburgh Sleep Quality Index in Australia Adults. Sleep and Biological Rhythms 2008; 222–227.

7. Cole J.C.,Motivala S. J., Buysse D.J. Validation of a 3-factor Scoring Model for the Pittsburgh Sleep Quality Index in Older Adults. Sleep 2006; 112–116.

8. Burkhalter H., Sereika S.M., Engberg S., others. Structure Validity of the Pittsburgh Sleep Quality Index in Renal Transplant Recipients: a Confirmatory Factor Analysis. Sleep and Biological Rhythms 2010; 274–281.

9. Giuseppe Curcio, Tempesta Daniela, Scarlata Simone, others. Validity of the Italian Version of the Pittsburgh Sleep Quality Index (PSQI). Neurological Sciences 2013.

10. Naser M.G.M., Parisa N., Zohreh O., others. The Reliability and Validity of the Persian Version of Pittsburgh Sleep Quality Index in Iranian People. Avicenna Journal of Neuro Psycho Physiology 2017.

11. Dunleavy Gerard, Bajpai Ram, Comiran Tonon Andre, others. Examining the Factor Structure of the Pittsburgh Sleep Quality Index in a Multi-Ethnic Working Population in Singapore. International Journal of Environmental Research and Public Health 2019.

12. Manzar, Md. Dilshad, others. Validity of the Pittsburgh Sleep Quality Index in Indian University Students. Oman Medical Journal 2015; 193–202.

13. Panda S., others. Sleep-related Disorders among a Healthy Population in South India. Neurology India 2012; 68–74.

14. R.B. Kline Principles and Practice of Structural Equation Modeling. Guilford Press. New York, New York.; 2010.

15. Rosseel Y. lavaan: An R Package for Structural Equation Modeling. Journal of Statistical Software 2012; 1–36.

16. Hirschfeld Gerrit, von Brachel Ruth Group Confirmatory Factor Analysis in R. Research & Evaluation 2014.

17. Audigier V., Husson F., Josse J. Cutoff Criteria for Fit Indexes in Covariance Structure Analysis: Cnventional Criteria versus New Alternatives. Structural Equation Model 1999; 1–55.

18. Panda S., others. A Principal Components Method to Impute Missing Values for Mixed Data. Advanced Data Analysis and Classification 2016.

19. Little Roderick. Calibrated Bayes, for Statistics in General, and Missing Data in Particular. Statistical Science 2011.

20. Enders Craig K., Bandalos Deborah L. The Relative Performance of Full Information Maximum Likelihood Estimation for Missing Data in Structural Equation Modelsata. Structural Equation Modeling: A Multidisciplinary Journal 2001.

21. Hallgren Kevin A., Witkiewitz Katie. Missing Data in Alcohol Clinical Trials: a Comparison of Methods. Alcoholism: Clinical and Experimental Research 2013.

